# Effects of Tocilizumab on Mortality in Hospitalized Patients with COVID-19: A Multicenter Cohort Study

**DOI:** 10.1101/2020.06.08.20125245

**Authors:** Javier Martínez-Sanz, Alfonso Muriel, Raquel Ron, Sabina Herrera, José A. Pérez-Molina, Santiago Moreno, Sergio Serrano-Villar

**Author notes:** Correspondence to: Javier Martínez-Sanz, MD, PhD. Department of Infectious Diseases, Hospital Universitario Ramón y Cajal, Facultad de Medicina, Universidad de Alcalá (IRYCIS). Carretera de Colmenar Viejo, Km 9.100, 28034 Madrid, Spain., Sergio Serrano-Villar, MD, PhD. Department of Infectious Diseases, Hospital Universitario Ramón y Cajal, Facultad de Medicina, Universidad de Alcalá (IRYCIS). Carretera de Colmenar Viejo, Km 9.100, 28034 Madrid, Spain. Co-senior authors (Santiago Moreno, Sergio Serrano-Villar).

## Abstract

**Background:** While there are no treatments with proven efficacy for patients with severe coronavirus disease 2019 (COVID-19), tocilizumab has been proposed as a candidate therapy, especially among patients with higher systemic inflammation.

**Methods:** We conducted a cohort study of patients hospitalized with COVID-19 in Spain. The primary outcome was time to death and the secondary outcome time to intensive care unit admission (ICU) or death. We used inverse-probability weighting to fit marginal structural models adjusted for time-varying covariates to determine the causal relationship between tocilizumab use and the outcomes.

**Results:** A total of 1,229 and 10,673 person/days were analyzed. In the adjusted marginal structural models, a significant interaction between tocilizumab use and high C-reactive protein (CRP) levels was detected. Tocilizumab was associated with decreased risk of death (aHR 0.34, 95% CI 0.16–0.72, p=0.005) and ICU admission or death (aHR 0.38, 95% CI 0.19–0.81, p=0.011) among patients with baseline CRP >150 mg/L, but not among those with CRP ≤150 mg/L. Exploratory subgroup analyses yielded point estimates that were consistent with these findings.

**Conclusions:** In this large observational study, tocilizumab was associated with a lower risk of death or ICU or death in patients with higher CRP levels. While the results of ongoing clinical trials of tocilizumab in patients with COVID-19 will be important to establish its safety and efficacy, our findings have implications for the design of future clinical trials and support the use of tocilizumab among subjects with higher CRP levels.

## Background

There are still no treatments with proven efficacy to prevent mortality in patients with severe coronavirus disease 2019 (COVID-19) pneumonia. However, various medications such as hydroxychloroquine, azithromycin, and lopinavir/ritonavir have been used off-label worldwide to minimize the impact of the current SARS-CoV-2 pandemic.^1^ Tocilizumab is an FDA-approved humanized monoclonal antibody against the soluble interleukin-6 (IL-6) receptor. It is widely used in the treatment of autoimmune disorders such as rheumatoid arthritis or cytokine release syndrome.^2,3^ Tocilizumab has been suggested as an effective treatment for severe COVID-19 pneumonia due to the increased interleukin 6 (IL-6) blood levels in patients with COVID-19^4^ and its correlation with a more severe lung damage.^5^ Tocilizumab is not currently approved for use by the FDA in COVID-19 patients. No efficacy results from observational studies or clinical trials in this disease have been published, and the available data comes from small studies with surrogate endpoints that are underpowered to detect significant clinical effects or lack a control group.^6–10^ Despite this absence of information, tocilizumab has been widely used due to its potential effect in the treatment of SARS-CoV-2-induced cytokine release syndrome in which IL-6 plays an important role.^5,11,12^

IL-6 determination is rarely available in clinical settings. However, C-reactive protein (CRP)—an inflammatory biomarker upstream in the IL-6 pathway—, is commonly used to monitor the activity of inflammatory diseases.^13^ In attempt to recruit the population with COVID19 with a higher probability to respond, some ongoing clinical trials of tocilizumab have considered heightened high CRP levels as an inclusion criterion (clinicaltrials.gov: NCT04346355 and NCT04356937).

Given the urgent need to respond to the COVID-19 pandemic, observational studies are important to evaluate clinical outcomes associated with the medications empirically used to treat COVID-19. However, critical analytical issues, including the risk of immortal time bias and indication bias from time-varying confounding^14^, challenge the validity of observational data in this setting.

Here, we investigate the association between tocilizumab use and mortality in a large cohort of hospitalized COVID-19 patients in Spain. We hypothesized that tocilizumab use would be associated with a lower risk of death and influenced by baseline systemic inflammation levels. We used marginal structural modeling to account for baseline and time-varying confounders.

## Methods

### Study design, setting, and data sources

We analyzed data from 2,047 subjects included in the *HM Hospitales* cohort—a multicenter cohort of patients admitted to any of the 17 hospitals in the HM Group in Madrid and diagnosed with COVID-19 from January 31^st^ to April 23^rd^, 2020. *HM Hospitales* made their anonymous dataset freely accessible to the international medical and scientific community. The dataset includes all available clinical information on patients diagnosed with COVID-19, confirmed by polymerase chain reaction in nasopharyngeal swabs or another valid respiratory sample. The dataset collects the different interactions in the COVID-19 treatment process including detailed information on diagnoses, treatments, admissions, intensive care unit (ICU) admissions, diagnostic imaging tests, laboratory results, discharge or death, and diagnostic and procedural records coded according to the International Statistical Classification of Disease and Related Health Problems (ICD-10) classification.^15^ We excluded patients younger than 18 years and those who died or were transferred to another facility within 24 hours after admission to the emergency department. This study was approved by the Ethics Committee at University Hospital Ramón y Cajal (, approval number 191/20).

### Endpoint

The primary end point was the time from study baseline to death. The secondary outcome was a composite event including admission to the ICU or death (hereafter ICU/death). Study baseline was defined as the first day of hospitalization.

### Statistical analysis

We tested the associations among the preadmission variables with treatment variable by Chi-square tests for categorical variables and Wilcoxon rank sum tests for continuous variables. We calculated the incidence rates of death and ICU/death and compared the time of death or the composite endpoint according to tocilizumab using Kaplan-Meier methods and log-rank tests. We fitted marginal structural models to estimate discrete time hazards of death according to tocilizumab use via an inverse probability treatment weight (IPTW) estimation to account for the non-randomized treatment administration of tocilizumab, baseline confounding, and time-varying confounders.^14,16^

We assumed that once a patient received tocilizumab, they remained on it until the end of follow-up. This assumption helped obtain a conservative estimate of the treatment hazard ratio analogous to intention-to-treat analysis in an unblinded randomized controlled trial. We structured the data set to allow for exposure, outcomes, right-censoring, and time-dependent covariates to change daily after admission. Propensity score logistic models predicted exposure at baseline and censoring over time as a result of recognized confounders of severe COVID-19^17,18^ including age, gender, comorbidities (hypertension, diabetes, ischemic heart disease, kidney disease, congestive heart failure, lung disease), oxygen blood saturation and need for oxygen therapy at baseline, and time-varying parameters of clinical severity (blood pressure, heart rate, total lymphocyte and neutrophil count, lactate dehydrogenase, alanine aminotransferase, urea, D-dimers, and CRP).

There were 1,229 subjects in the dataset with the information needed to fit marginal structural models. The characteristics of the individuals not included due to missing data in the information required for the statistical modelling strategy are shown in **Table S1** in the Supplement. The main differences between groups in the analyzed population were comparable to those found in the population with missing information. IPTW were stabilized and truncated below the first percentile and above the 99th percentile. The models included a main term for the exposure and a flexible functional form of time, that is, restricted cubic splines with 5 knots set at the first, 25^th^, 50^th^, 75^th^, and 99^th^ percentiles of the subjects’ day of follow-up, and the interaction term between tocilizumab and elevated CRP levels (>150 mg/L, cut- off selected based on the 75^th^ percentile value, 143 mg/L). The interaction term between tocilizumab and CRP was significant, and thus we report the adjusted (weighted) hazard ratios (HRs) derived from marginal structural models for the primary and secondary outcomes segregated by CRP levels.

We planned exploratory sensitivity analyses restricted to subjects who received specific concomitant treatments against SARS-CoV-2 (corticosteroids, hydroxychloroquine, azithromycin, lopinavir/ritonavir). Due to the recognized prognostic value of lymphocyte counts and D-dimer levels, we also performed sensitivity analysis to explore the possible confounding effect of D-dimer >1000 ng/mL (upper limit of the normal range in the reference laboratory) or absolute lymphocyte count <1000/uL (lower limit of the normal range in our reference laboratory). Statistical analyses were performed using Stata v. 16.0 (StataCorp LP College Station, TX, USA). Figures 1 and 2 were generated using Prism v.8.0 (GraphPad, La Jolla, California).

**Figure 1.**
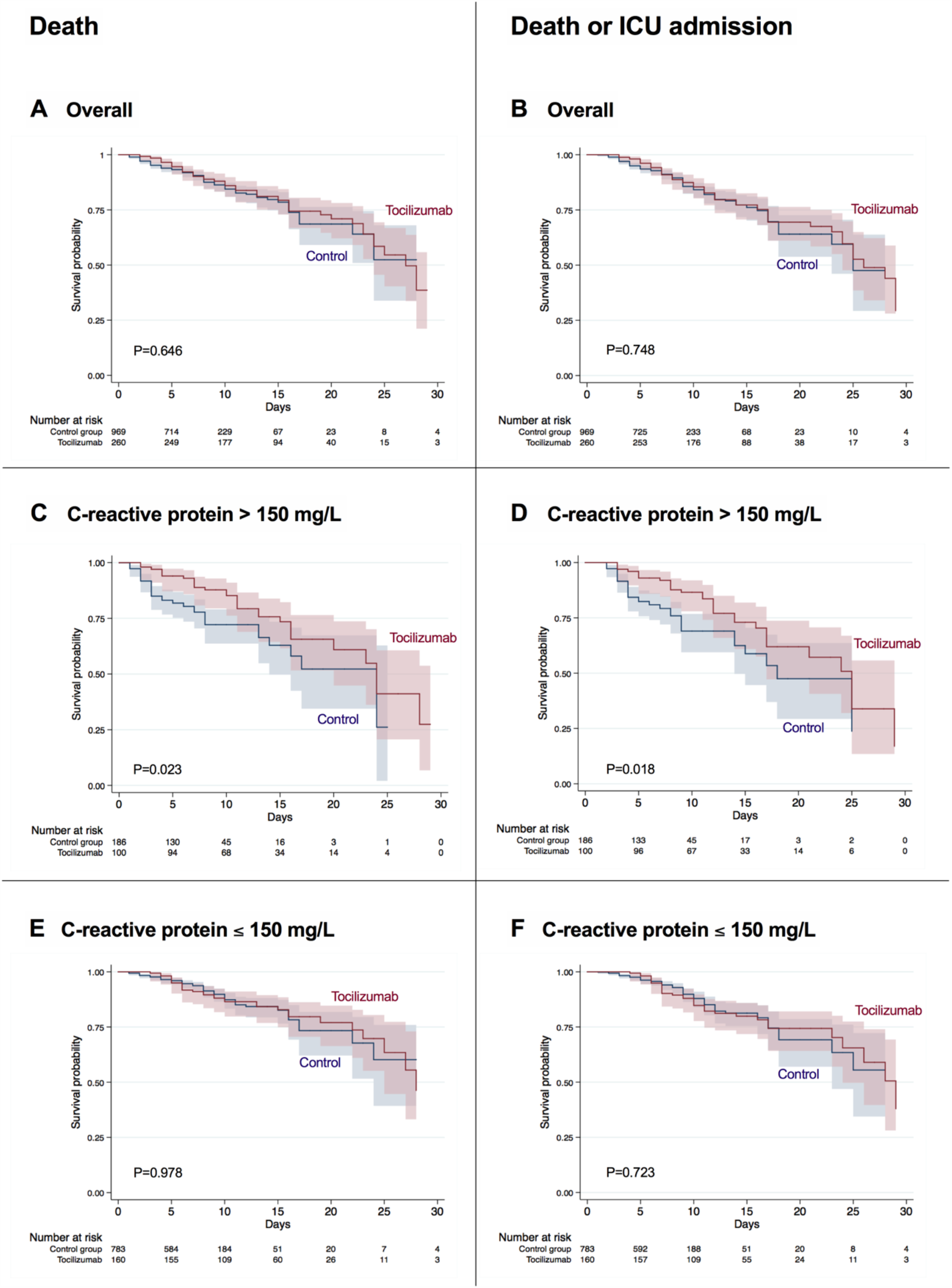
Kaplan-Meier failure estimates for death and ICU/death.

**Figure 2.**
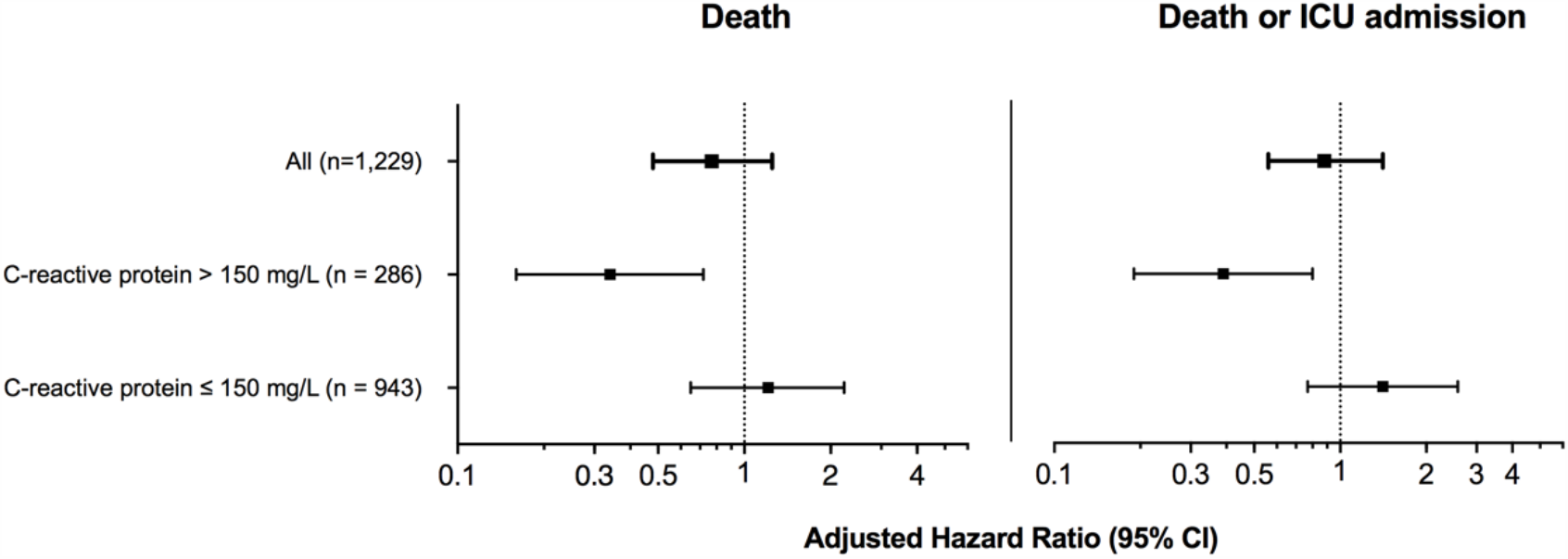
Adjusted hazard ratios for primary and secondary endpoints. Weighted hazard ratios derived from marginal structural models adjusted for sex, age, comorbidities (hypertension, diabetes, ischemic heart disease, chronic kidney disease, congestive heart failure, lung disease), need for oxygen therapy at baseline, oxygen blood saturation, and time-varying parameters of severity (blood pressure, heart rate, total lymphocyte and neutrophil count, LDH, ALT, urea, D-dimer, and CRP). *Abbreviations: ALT, alanine aminotransferase; CI, confidence interval; ICU, intensive care unit; LDH, lactate dehydrogenase*.

## Results

### Study population

We analyzed 1,229 subjects accounting for 11,900 observations and 10,673 person-days of follow-up who were diagnosed with COVID-19 in HM hospitals between January 31^st^ and April 23^rd^, 2020 and have the information needed for IPTW estimation. We excluded 99 patients because they died, were discharged, or were transferred to a different hospital within 24 hours after admission to the emergency department. A total of 181 subjects (14.7%) died, 82 (6.7%) were admitted to the ICU, and 186 had a composite outcome of death or ICU admission (15.1%). The median time to censoring date was 6 (IQR 3-9) days. Of the 1,229 patients, 260 (21%) received a median total dose of 600 mg (IQR 600–800 mg) of tocilizumab. The first dose was administered at a median time of 4 (IQR 3 – 5) days from inpatient admission. The distribution of the patient’s characteristics according to tocilizumab use is shown in **Table 1**. Compared to the control group, there was a higher frequency of men and previous lung disease in the tocilizumab group while controls were significantly older and had a higher prevalence of diabetes. As expected, there were small differences between both groups in some of the baseline vital signs and laboratory parameters that were indicative of greater disease severity in the tocilizumab group than in the control group.

**Table 1.** Population baseline characteristics according to the use of tocilizumab.

|  | **Control group**<br>**(n = 969)** | **Tocilizumab**<br>**(n = 260)** | **p** |
| --- | --- | --- | --- |
| **Age, median (IQR)** | 68 (57 - 80) | 65 (55 - 76) | 0.017 |
| **Gender, n (%)** |  |  |  |
| Male | 574 (59) | 191 (73) | <0.001 |
| Female | 395 (41) | 69 (27) |  |
| **Past medical history, n (%)** |  |  |  |
| Hypertension | 227 (23) | 44 (17) | 0.025 |
| Diabetes | 233 (24) | 47 (18) | 0.042 |
| Congestive heart failure | 31 (3) | 5 (2) | 0.279 |
| Coronary artery disease | 77 (8) | 21 (8) | 0.945 |
| Chronic kidney disease | 53 (5) | 11 (4) | 0.425 |
| Lung disease | 94 (10) | 39 (15) | 0.015 |
| **Vital signs at admission** |  |  |  |
| Heart rate (bpm), median (IQR) | 83 (72 - 94) | 84 (75 - 96) | 0.059 |
| Systolic blood pressure (mmHg), median (IQR) | 127 (114 - 140) | 128 (115 - 141) | 0.592 |
| Temperature (°C), median (IQR) | 36.5 (36.0 - 37.1) | 36.8 (36.1 - 37.4) | 0.003 |
| Peripheral oxygen saturation (%), median (IQR) | 94 (92 - 96) | 91 (86 - 94) | <0.001 |
| Needing supplemental oxygen | 241 (25) | 54 (21) | 0.169 |
| **Baseline laboratory** |  |  |  |
| Absolute lymphocyte count (cells/μl), median (IQR) | 1050 (770 - 1440) | 890 (630 - 1225) | <0.001 |
| Absolute neutrophil count (cells/μl), median (IQR) | 4670 (3350 - 6560) | 5425 (3710 - 8165) | <0.001 |
| LDH (U/L), median (IQR) | 523 (408 - 664) | 669 (566 - 829) | <0.001 |
| ALT (U/L), median (IQR) | 26 (16 - 45) | 32 (21 - 53) | <0.001 |
| Urea (mg/dL), median (IQR) | 33 (25 - 48) | 22 (26 - 46) | 0.957 |
| C-reactive protein (mg/L), median (IQR) | 64 (27 - 122) | 113 (64 - 220) | <0.001 |
| D-dimer (ng/mL), median (IQR) | 720 (443 - 1389) | 809 (519 - 1270) | 0.127 |
| Interleukin 6 (pg/mL), median (IQR) | 27 (7 - 60) | 70 (26 - 182) | <0.001 |
| **Outcome** |  |  |  |
| Non-ICU length of stay (days), median (IQR) | 8 (5 - 10) | 13 (10 - 18) | <0.001 |
| ICU admission, n (%) | 32 (3) | 50 (19) | <0.001 |
| ICU length of stay (days), median (IQR) | 2 (1 - 3) | 6 (2 - 11) | 0.002 |
| Mortality, n (%) | 120 (12) | 61 (23) | <0.001 |
| ICU or mortality, n (%) | 120 (12) | 66 (25) | <0.001 |

All variables were available in the 1,229 subjects, with exception of IL-6, which was measured only in 88 individuals.
Abbreviations: ALT, alanine aminotransferase; ICU, intensive care unit; IQR, interquartile range; LDH, lactate dehydrogenase.

### Primary and secondary endpoints

The 1,229 subjects accounted for 11,900 observations, and the crude incidence rate of death was 17.8 (95% CI, 13.8 - 22.8) per 1000 persons-days in the tocilizumab group vs. 16.6 (95% CI, 13.9 - 19.8) per 1000 persons-days in the control group, yielding an incidence rate ratio of 1.07 (95% CI, 0.77 - 1.47). In the composite outcome of ICU/death, the crude incidence rate was 19.3 (95% CI, 15.1 - 24.5) in the tocilizumab group vs. 16.3 (95% CI, 13.6 - 19.5) per 1000 persons-days in the control group yielding an incidence rate ratio of 1.18 (95% CI, 0.86 – 1.61).

We used Kaplan-Meier estimates as a first approach to visualize the cumulative probabilities of death and ICU/death. We did not observe differences in the estimates of death or ICU/death in the pooled analysis (**Figure 1A-B**). However, we found a significantly lower cumulative probabilities of both outcomes among patients with baseline CRP levels above >150 mg/L (**Figure 1C-D**) but not among those with CRP ≤150 mg/L (**Figure 1E-F)**.

In the unadjusted marginal structural model analysis, tocilizumab was associated with a higher risk of death (HR 1.53, 95% CI 1.20 – 1.96, p=0.001) and ICU/death (HR 1.77, 95% CI 1.41 – 2.22, p<0.001). However, this effect disappeared in the adjusted analyses in which we found a significant interaction between tocilizumab use and CRP values (p=0.023 and p=0.012 for primary and secondary endpoints, respectively). Subjects who received tocilizumab and had baseline CRP levels above 150 mg/L experienced lower rates of death (aHR 0.34, 95% CI 0.17 - 0.71, p=0.005) and ICU admission/death (aHR 0.39, 95% CI 0.19 – 0.80, p=0.011) than those who did not receive tocilizumab. This effect was not observed among patients with baseline CRP levels ≤150 mg/dL (**Figure 2**).

**Figure 3 and Table S2** show the adjusted hazard ratios for exploratory sensitivity analyses restricted to patients with baseline lymphocyte count <1000 cell/µl and baseline D-dimer >1000 ng/mL segregated by CRP levels. The results are consistent with the principal analysis. Individuals with baseline CRP levels higher than 150 mg/dL maintained a lower risk of death and ICU/death, but no significant effects of tocilizumab were found among those with low CRP levels.

**Figure 3.**
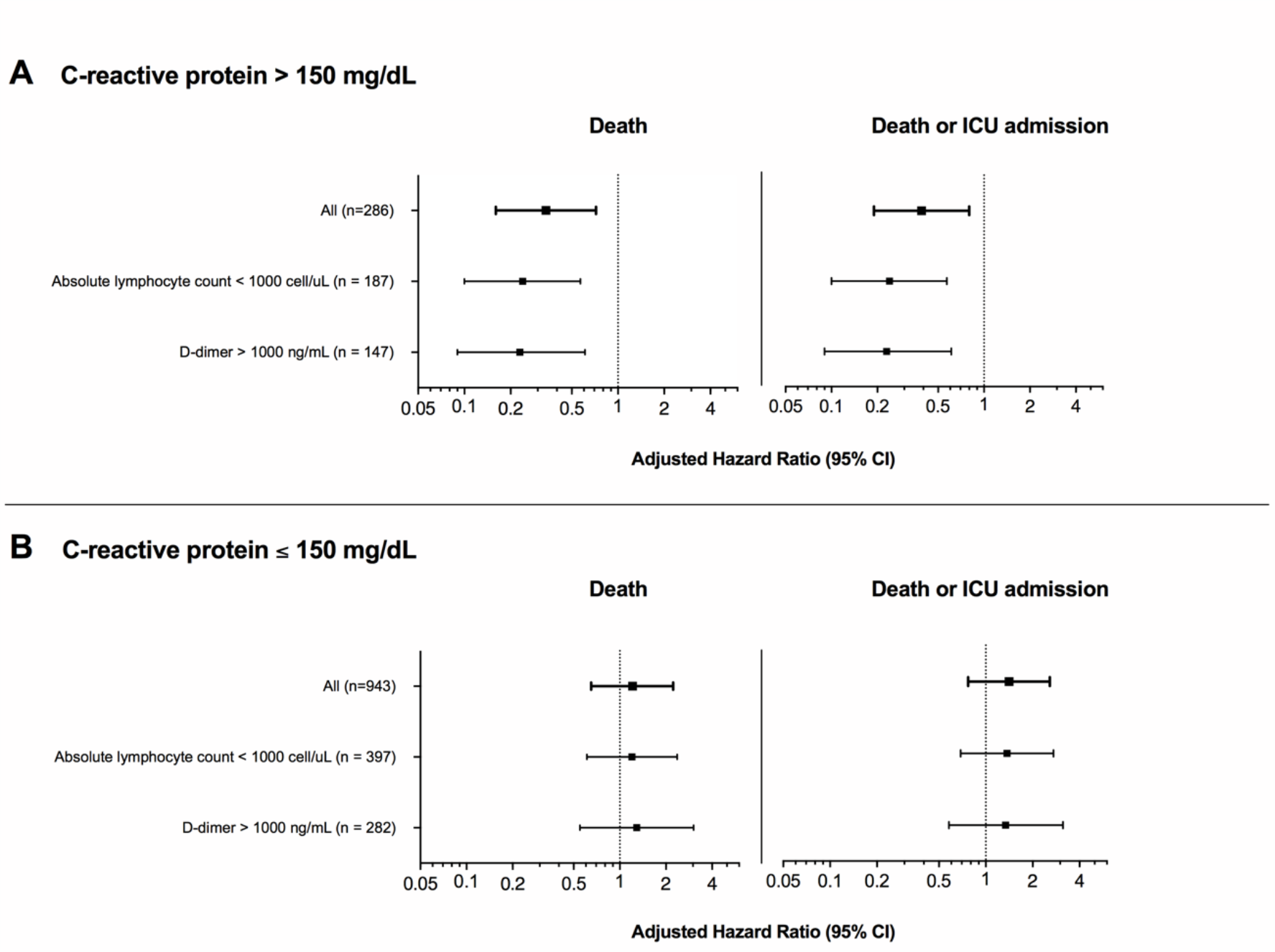
Adjusted hazard ratios for sensitivity analyses according to baseline laboratory absolute lymphocyte counts and D-dimers. Weighted hazard ratios derived from marginal structural models adjusted for sex, age, comorbidities (hypertension, diabetes, ischemic heart disease, chronic kidney disease, congestive heart failure, lung disease), need for oxygen therapy at baseline, oxygen blood saturation, and time-varying parameters of severity (blood pressure, heart rate, total lymphocyte and neutrophil count, LDH, ALT, urea, D-dimer, and CRP). *Abbreviations: ALT, alanine aminotransferase; CI, confidence interval; ICU, intensive care unit; LDH, lactate dehydrogenase*.

We also explored the effects of concomitant therapies against SARS-COV-2 in sensitivity analyses restricted to subjects who received corticosteroids (n=582), hydroxychloroquine (n=1,134), azithromycin (n=812), or lopinavir/ritonavir (n=753) (**Table S2**). The effect sizes among subjects with baseline CRP >150 mg/L were very similar to those observed in the principal analyses for both the primary and secondary outcomes (all p-values <0.05 except azithromycin and lopinavir/ritonavir, with p=0.105 in the primary and p=0.079 in the secondary outcome, respectively).

## Discussion

This is the first study to evaluate the effects of tocilizumab on the mortality of hospitalized patients with COVID-19. While the overall risk of death or ICU admission did not differ among patients who received tocilizumab to those who did not and had CRP levels ≤150 mg/L, we found a 66% reduction in the risk of mortality in subjects with baseline CRP levels >150 mg/L who received tocilizumab. Exploratory sensitivity analyses to account for the use of concomitant therapies against SARS-CoV-2 or the presence of other analytical parameters predictive of poor prognosis yielded consistent treatment effect estimates.

Early reports suggest the systemic inhibition of IL-6 signaling to treat COVID-19. Both SARS-CoV-2^19^ and SARS-CoV^20^ exploit angiotensin converting enzyme II (ACE2) to infect pulmonary and intestinal epithelial cells and activate NF-kappa B upregulation of proinflammatory cytokines^21^ such as IL-6, IL-8, and gamma interferon-inducible protein 10, which stimulates monocyte-derived and dendritic cells. While causation has yet to be confirmed, excessive pulmonary and systemic cytokine release sustained in part by IL-6 signaling is a candidate mechanism explaining the infiltration of activated neutrophils into the alveolar space and subsequent diffuse lung damage found in patients with severe COVID-19.^22^ The original observational study of 21 patients with COVID-19 in Wuhan reported that tocilizumab resulted in resolution of fever in all patients one day after treatment and improvement of peripheral oxygen saturation and lymphopenia. Moreover, 19 of 20 patients who received tocilizumab presented a radiological improvement in the CT scan after 5 days.^7^ In a single-arm study of 63 patients with a pro-inflammatory and pro-thrombotic state due to severe COVID-19, treatment with tocilizumab was associated with a decrease in CRP, D-dimer, and ferritin levels.^6^ Thus, tocilizumab and other repurposed medications have been widely used off-label to treat COVID-19 in an attempt to mitigate the dramatic clinical consequences of SARS-Cov-2 pandemic, despite the lack of information on the effects of tocilizumab on robust clinical outcomes.

The first confirmed case of COVID-19 in Spain occurred on January 31 of 2020, and 235,772 cases had been identified by May 25^th^, 2020.^14^ Due to the impact of the disease at a time when randomized trials were lacking, many protocols were developed, and the use of 2-3 doses of tocilizumab adjusted by weight was allowed by the Spanish National Guidelines as a possible treatment for COVID-19.^23^ Shortly after the release of this document, the number of doses was restricted to a single dose adjusted by weight (400 mg or 600 mg) to avoid a tocilizumab shortage.

In this multicenter cohort, we could compare 260 subjects who received tocilizumab with 969 who did not. These subjects accounted for 181 deaths, 82 ICU admissions, and 186 combined events of ICU admission or mortality. We selected an analytical approach capable of dealing with the potential confounders inherent to observational studies in which subjects receiving tocilizumab were expected to have more risk factors for clinical progression and greater disease severity at baseline. In our cohort, controls were significantly older and had a higher prevalence of hypertension, which are the risk factors that have been more robustly associated with severe COVID-19 and death.^4,17,24,25^ However, subjects who received tocilizumab tended to have a greater prevalence of other potential risk factors for disease severity such as lung disease, as well as differences in baseline vital signs and laboratory parameters indicative of greater disease severity. All of these factors were included as covariates, and the estimates were consistent across the two endpoints analyzed. We found a strong and consistent protective effect of tocilizumab among patients with CRP levels above 150 mg/L (aHR 0.34, 95% CI 0.17 – 0.71).

The selection of the modeling strategy was a critical decision. Longitudinal studies in which exposures, confounders, and outcomes are measured repeatedly over time can facilitate causal inferences about the effects of exposure on outcome.^14^ However, there are key analytical issues in this setting, including the risk of immortal time bias (i.e., the requirement for patients to survive long enough to receive the intervention of interest, which can lead to a potentially incorrect estimation of a positive treatment effect), and indication bias from time-varying confounding (e.g., the use of tocilizumab following elevations of CRP).^14^ Standard regression models for the analysis of cohort studies with time-updated measurements may result in biased estimates of treatment effects if time-dependent confounders affected by prior treatment are present.^14,26^ Marginal structural models are a powerful method for confounding control in longitudinal study designs that collect time-varying information on exposure, outcome, and other covariates, such as the present one.^14,16^

Our study has a number of limitations. As with any observational study, there is still a risk of unmeasured confounding. Tocilizumab targets the IL-6 receptor, and thus using baseline IL-6 levels instead of CRP in the interaction term with tocilizumab use could have helped to better discriminate the population benefiting most from tocilizumab treatment. Although IL-6 measurements are rarely available in clinical settings, CRP is widely accessible and is an inflammatory biomarker upstream of the IL-6 pathway.^13^ Hence, we doubt that the use of CRP instead of IL-6 limited the scope of the results. Ongoing trials of tocilizumab in COVID-19 have also considered heightened CRP instead of IL-6 to identify patients with heightened inflammation and, therefore, potential greater benefit with this treatment (clinicaltrials.gov: NCT04346355 and NCT04356937). In addition, the results should be interpreted cautiously and must not be taken as confirmatory of tocilizumab efficacy because of the relatively wide confidence intervals in the principal analysis. The sensitivity analyses suggested that patients with high CRP and high D-dimer levels or lymphopenia may also be target populations for tocilizumab use. However, despite the fact that the size effects observed here were consistent with those obtained in the principal analyses, the sub-analyses must only be interpreted as exploratory and hypothesis-generating.

The main strengths of the study include the large sample size and multicenter contribution that is representative of a real-life setting. This can allow generalizability of the results. The availability of daily information on covariates defining treatments and laboratory parameters allowed us to control confounding issues using marginal structural models. Finally, the high number of outcomes powered the statistical analysis, and the results are novel and biologically plausible. In summary, we analyzed a large number of consecutive patients hospitalized with COVID-19 and found that tocilizumab was associated with a lower risk of mortality or ICU admission/mortality among patients with CRP >150 mg/L, but not among those with lower CRP levels. Although the results of ongoing clinical trials of tocilizumab in patients with COVID-19 are mandatory to establish its safety and efficacy, our findings have implications for the design of future clinical trials and support the use of tocilizumab among subjects with higher levels of inflammatory markers.

## Data Availability

The dataset is freely available upon request for a research project to

## Acknowledgments

We acknowledge all study participants who made this research possible. We thank *HM Hospitales* group for releasing the dataset used to perform this research to the scientific community.

